# A Pathology-Trained Lipid Aging Clock Reveals Accelerated Aging and Prognostic Sphingolipid Signatures in Pancreatic Disease

**DOI:** 10.1101/2024.09.03.24311998

**Authors:** Maximilian Unfried, Amaury Cazenave-Gassiot, Evelyne Bischof, Michal Holcapek, Morten Scheibye-Knudsen, Markus R. Wenk, Jan Gruber, Brian K. Kennedy

## Abstract

Altered lipid metabolism is increasingly recognized as a feature of aging and age-related disease, including cancer. Here, we present a lipid-based biological aging clock developed from serum lipidomics data of 443 patients with pancreatic ductal adenocarcinoma (PDAC). Using penalized Cox proportional hazards models, we derived a risk-equivalent Lipid Age and Lipid Age Acceleration (LAA) metric and evaluated its relationship to survival, healthy aging, and pancreatitis. Despite being trained exclusively on a pathological PDAC cohort, the lipid clock predicted chronological age in healthy individuals with high accuracy (Pearson r=0.86; median absolute error = 3.46 years), indicating that pathological cohorts can retain biologically meaningful aging signals. Both PDAC patients and, in exploratory analyses, pancreatitis patients exhibited accelerated lipid aging relative to healthy individuals, with mean LAA values of 5.57 and 2.71 years, respectively. Increased continuous LAA was significantly associated with worse overall and progression-free survival and remained predictive of overall mortality even after adjusting for standard clinical cancer measures, staging, and the conventional tumor marker CA19-9 (HR=1.09 per 1-year increase, p<0.005). Specific individual lipid species, particularly sphingolipids, carried independent prognostic value. Lipid age acceleration was associated with pancreatic disease and with lipidomic patterns consistent with systemic aging-associated and inflammatory remodeling independent of traditional oncological risk parameters. Because the model was developed and evaluated within a single parent dataset and analytical platform, external validation in independent cohorts will be needed to establish broader generalizability.

## Introduction

Aging and age-related diseases are major contributors to the global disease burden. Measuring the divergence between biological and chronological age, and the change in mortality risk in patients with chronic illness is therefore of considerable interest. While most aging clocks have been developed in healthy cohorts, clocks that inform disease staging, progression, and prognosis are needed in clinical practice.

Lipids play a central role in the pathology of aging and associated disorders. While thousands of lipid species exist in human serum, routine lipidomic approaches typically quantify only a few hundred lipids(1–3). The abundances of many lipid species undergo dynamic changes with disease, as well as during tissue and organismal aging(4,5), with some multi-omics studies finding that lipids comprise up to 40% of the metabolites changing with age(6). Dysregulation of lipid metabolism is observed in various age-related conditions, including cardiovascular and neurodegenerative diseases, Diabetes mellitus, and various forms of cancer(7–11). Lipid species such as cholesterol, ceramides, sphingomyelins, and lysophosphatidylcholines are already used or proposed as biomarkers in several of these conditions(8,12–16).

However, deconvoluting signatures of intrinsic aging from disease-associated lipid changes remains challenging, and accurate lipid-based biomarkers of biological aging in humans are still lacking.

In aging biology and geromedicine, biological aging clocks have emerged as informative tools to quantify biological age at the individual and population level, including in clinical trials of longevity interventions(17–21).

Aging clocks based on machine learning and statistical approaches have been developed from methylation, glycans, proteins, metabolites, lipids, RNA(22–31), and clinical biomarkers(32,33),and can predict all-cause mortality, frailty, and chronic disease better than chronological age alone(22,34–36). While epigenetic clocks have achieved high predictive accuracy, one advantage of metabolite-based clocks is that age-dependent changes can often be interpreted mechanistically through specific biological pathways(37,38). Epigenetic age acceleration has also been associated with cancer risk and mortality(39–42).

Pancreatic ductal adenocarcinoma (PDAC) is one of the most severe cancers with a 5-year survival rate of less than 10%(43). Pooled analysis of three cohorts showed that Hannum, Horvath and PhenoAge intrinsic epigenetic age acceleration are significantly associated with pancreatic cancer risk, but not with pancreatic cancer survival(44). In PDAC, lipid metabolism is profoundly dysregulated, with recurrent alterations in sphingomyelins, ceramides, and phosphatidylcholines in patient serum, highlighting the potential for precise lipid-based biomarkers of the disease(15,45–47). While DNA methylation clocks primarily reflect cumulative aging burden and long-term biological aging processes, lipidomic clocks may capture a more dynamic picture of current physiological resilience by integrating signals related to inflammation, immune function, mitochondrial health, and cancer-associated metabolic dysregulation.

In this study, we show that the serum lipidome can be used to derive a risk-equivalent lipid aging signal associated with biological aging and disease prognosis. Building on our findings from lipid clocks in model organisms(28) and human post-mortem prefrontal cortex lipidomic data(48), we translate this concept to human serum and show a direct relationship between pancreatic disease and accelerated aging. We present a new human lipid age clock that predicts survival in PDAC patients and is linked to inflammatory lipid signals. We further show that aging clocks can be constructed from pathological cancer cohorts, highlighting the potential of large cancer datasets for aging clock research.

## Materials and Methods

### Dataset

The dataset used in this study was collected by Wolrab et al.(15) and consists of 830 serum samples gathered in a 3-phase biomarker discovery study. Specifically, the dataset used here originated from the Phase III (verification) of said study (see Supplementary for Material and Methods for more details on sample collection and processing). The phenotypes of the participants were healthy controls, patients with pancreatic ductal adenocarcinoma, or pancreatitis with specifics on demographics provided in Sup. Tab. 1. The dataset comprised 546 PDAC patients, of whom 443 had overall survival information and were included in survival analyses. A total of 202 lipid species were quantified and after filtering on a strict criterion to retain only species measured in all samples and for which fewer than 5% of samples shared the same exact value, 102 lipids were entered into the model (Sup. Tab. 2).

**Table 1:**
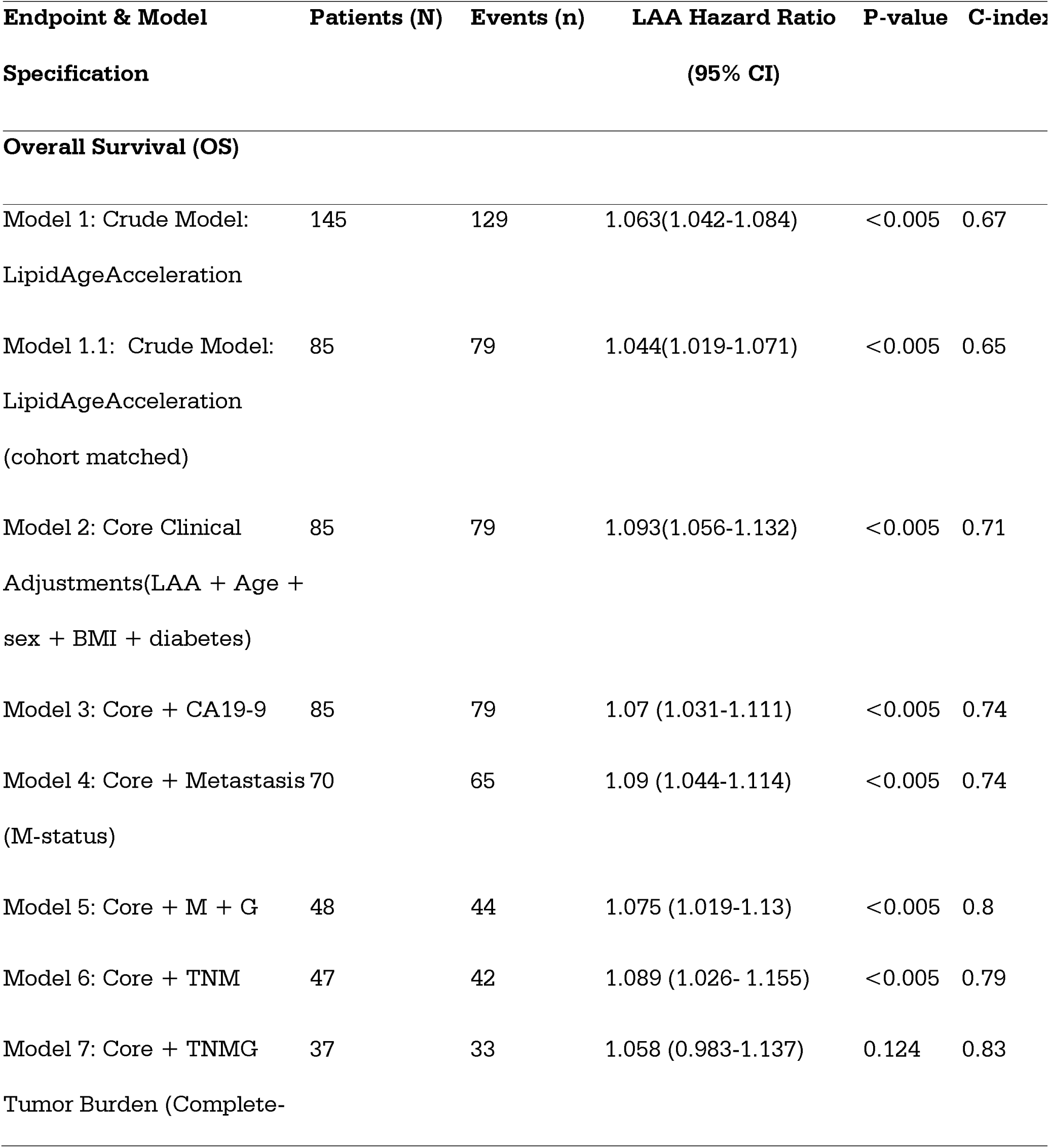
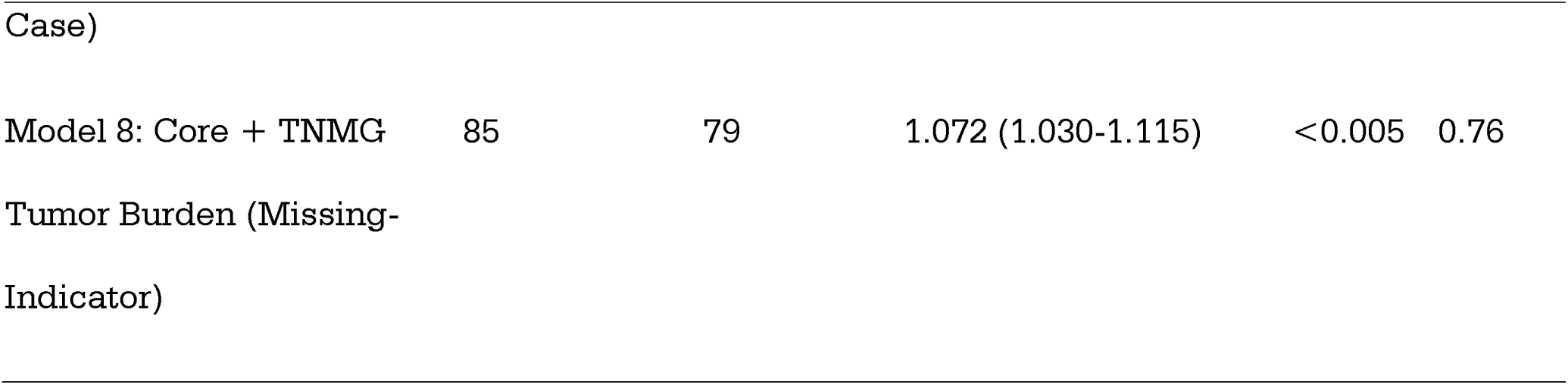
Multivariable Cox proportional hazards regression models for Overall Survival (OS) based on Lipid Age Acceleration (LAA). The table displays patient numbers, mortality events, Hazard Ratios (HRs) with 95% Confidence Intervals (CIs), P-values, and Concordance Index (C-index) across sequentially adjusted models. Model 1 assesses the unadjusted (crude) effect of LAA in the full cohort, while Model 1.1 evaluates the crude effect within the cohort-matched subset. Model 2 (Core) adjusts for baseline clinical covariates (chronological age, sex, BMI, and diabetes status). Subsequent models (Models 3–6) evaluate the independent prognostic value of LAA when accounting for clinical biomarkers (CA19-9) and tumor burden characteristics. Models 7 and 8 contrast handling strategies for missing staging data using complete-case analysis and the missing-indicator method, respectively. Incremental increases in the C-index demonstrate improved model discriminative capacity with advanced clinical stratification.

**Table 2:**
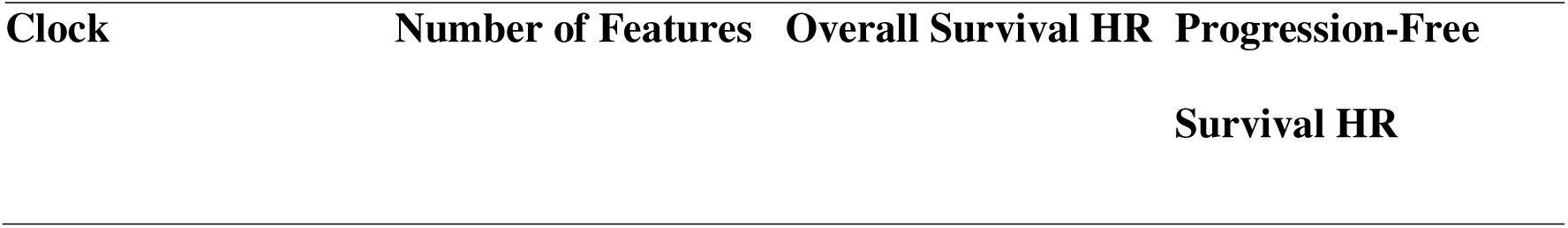
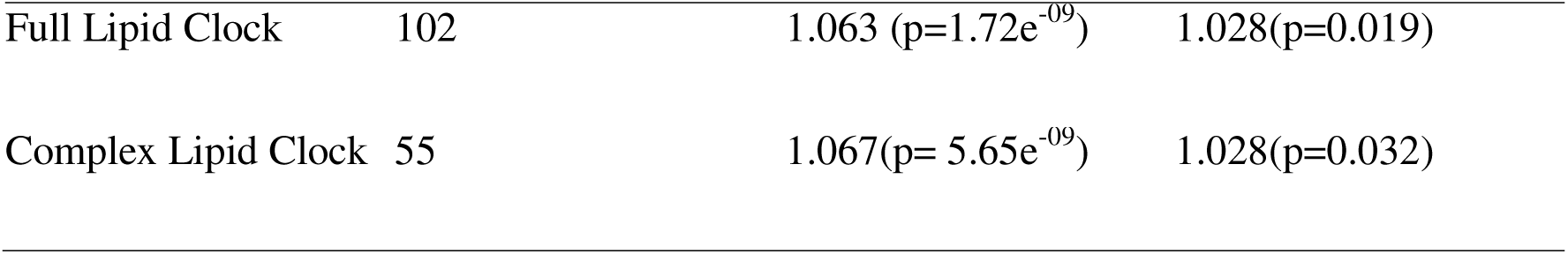
Methodological comparison of the Full Lipid Clock versus the streamlined Complex Lipid Clock framework for survival outcomes. The table details feature counts alongside baseline Hazard Ratios (HRs) and corresponding p-values for Overall Survival (OS) and Progression-Free Survival (PFS) modeling. Hazard ratios reflect the independent risk associated with each 1-year increase in Lipid Age Acceleration (LAA). The comparison demonstrates that reducing the biomarker architecture by approximately 46% (from 102 down to 55 complex lipid features) preserves prognostic power, maintaining highly consistent risk parameters and statistical significance profiles across both clinical oncology endpoints.

| Clock | Number of Features | Overall Survival HR | Progression-Free Survival HR |
| --- | --- | --- | --- |
| Full Lipid Clock | 102 | 1.063 (p=1.72e<sup>-09</sup>) | 1.028(p=0.019) |
| Complex Lipid Clock | 55 | 1.067(p= 5.65e<sup>-09</sup>) | 1.028(p=0.032) |

### Constructing the Lipid Clock and Model training

Similar to the PCAge clinical clock, the LipidClock consists of the combination of two penalized Cox proportional hazards models(35) that create a risk-equivalent age(49). The underlying Cox proportional hazard model is a regression model which regresses between the survival time of patients and one or more predictor variables(50). Through an Elastic Net penalty, the model selects which subset of features, in our case chronological age and lipid species, are associated with mortality, and with what kind of magnitude.

We optimize the parameters for the penalized cox proportional hazard model through grid search and stratified 5-fold cross validation. Details can be found in the supplementary materials.

The final lipid clock model is trained with 67% of the PDAC data, and 33% are held out for evaluation. We used a stratified train-test split to keep the distribution of age and overall survival events (Sup. Tab. 3). Moreover, we evaluate it against the healthy and pancreatitis samples. Details on parameter estimation and derivation of the algorithm are given in the supplementary materials.

We performed a sensitivity analysis to evaluate the effect of the mortality-doubling-time (MDT) parameter used to convert hazard-ratio differences into age-equivalent LipidAge and LAA units. The clock outputs were recalculated across a plausible range of MDT values (6–10 years), and for each scaling choice we reassessed age-prediction metrics, cohort-level LAA distributions, and univariable and multivariable survival associations. This analysis was designed to test whether the principal conclusions depended on the specific age-scaling constant rather than the underlying hazard structure.

## Results

### Construction of a PDAC-derived lipid aging signal

We developed a Lipid Aging Clock to predict mortality and biological risk-equivalent age in patients diagnosed with Pancreatic Ductal Adenocarcinoma, using two penalized Cox proportional hazards models – one trained on the lipid profile, and the other on solely chronological age, providing us with hazard ratios for individual lipids and chronological age. The final lipid model was ridge-like (L1-ratio=0.05), containing 102 non-zero lipid features. The age-only and lipid-based Cox models were combined to derive the Lipid Age Acceleration (LAA) metric, based on the idea of risk-equivalent age(49).

To test if the human serum lipidome can be used as an accurate predictor of biological age we trained a LipidClock on data from 294 individuals with pancreatic ductal adenocarcinoma and evaluated it on a holdout test set of 145 PDAC patients (Sup. Tab. 3), 22 patients with pancreatitis and 262 healthy individuals (Sup. Tab 2.). To date, most aging clocks have been generated from healthy populations, or at least not individuals with one condition. Our approach is a test of the hypothesis that reliable measure of aging can be extracted from a pathological cohort. If successful, this approach would open a much wider range of cohorts in which it is possible to determine biological age and enhance the capacity of this methodology to disentangle the role of aging in disease progression.

### LipidAge preserves age structure while PDAC and pancreatitis exhibit positive lipid age acceleration

The LipidClock successfully predicts the chronological age of healthy individuals (Fig. 1A), and we find that it can accurately predict the biological age acceleration in healthy individuals with a mean and median absolute error of 4.82 years and 3.46 years (Sup. Tab. 4).

**Figure 1:**
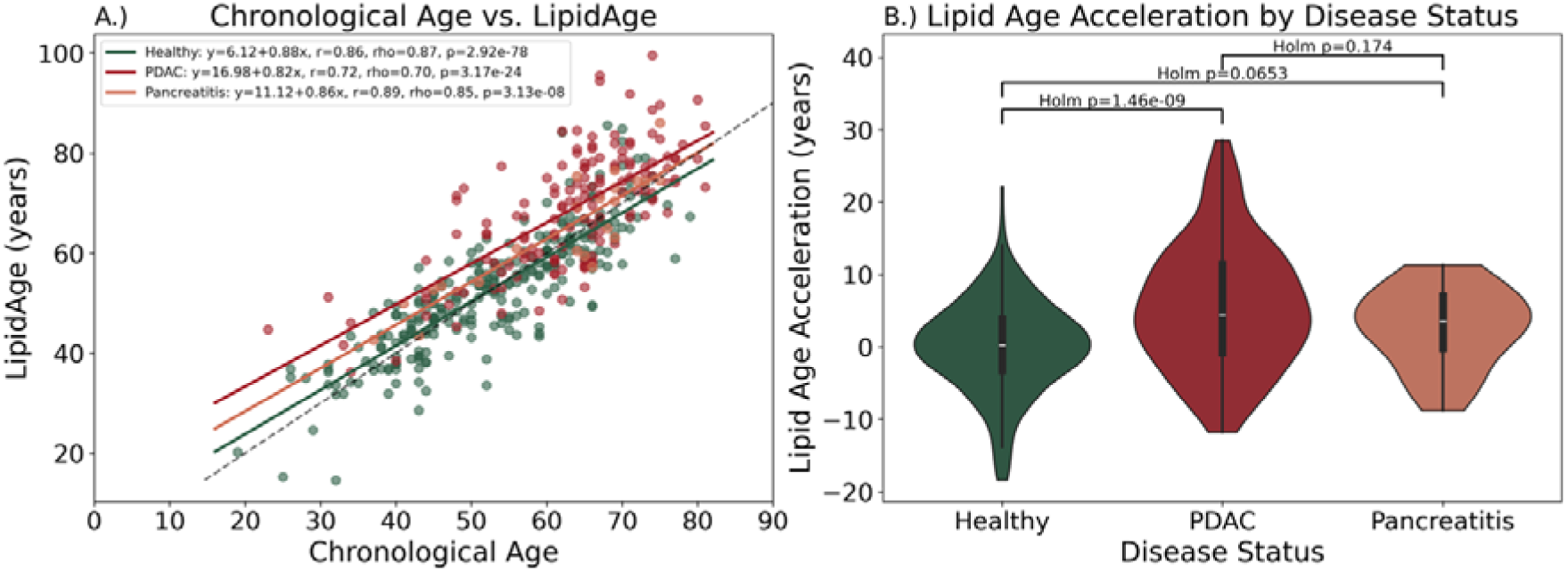
Evaluation of the lipidomics-based aging clock (LipidClock) and Lipid Age Acceleration (LAA) across healthy controls and pancreatic disease cohorts. A.) Correlation between chronological age and predicted LipidAge. Scatter plot demonstrating the linear relationship between chronological age (x-axis) and predicted LipidAge (y-axis) stratified by cohort: Healthy controls, PDAC, and Pancreatitis patients. Linear regression lines, Pearson correlation coefficients (r), Spearman’s rank correlation coefficients (]), and corresponding p-values are shown for each group, indicating robust model calibration. The dashed gray line represents the line of identity (y=x), where chronological age perfectly matches biological lipid age. B.) Quantification of Lipid Age Acceleration (LAA) by disease status. Violin plots illustrating the distribution and density of LAA (measured in years) across the three clinical cohorts. Pairwise group differences were evaluated using post-hoc analysis with Holm correction for multiple comparisons. Significant lipidomic age acceleration is observed in PDAC patients compared to healthy controls (p=1.46×10−9), while the exploratory pancreatitis analysis showed a marginal, non-significant trend toward acceleration (p=0.0653).

Given that the clock was constructed from serum of PDAC patients, we were surprised by its accuracy in forecasting aging in healthy individuals. Conversely, predicting the biological age of pancreatic cancer patients yields a mean and median absolute error of 8.09 and 6.8 years respectively, indicating that on a population level the lipid profile of PDAC patients might show more variation, yielding larger errors (Sup. Tab. 4). Accordingly, the Pearson correlation between chronological and LipidAge for healthy individuals is 0.86 (p=2.92e-78), 0.72(p=3.17e-24) for PDAC patients, and 0.89 (p=3.13e-08) for patients with pancreatitis (Fig. 1A). These data suggest that healthy people are more alike, whereas there are many different molecular variations and clinical phenotypes of a disease, especially when as complex as cancer.

Generally, when developing an aging clock on data of healthy individuals, the predicted biological age may be expected to be accelerated for various age-related diseases, especially cancer(22,39,41). Thus, we reasoned that the converse may also be true: developing an aging clock on diseased individuals should predict decelerated age for healthy individuals. This is indeed the case with PDAC patients, having an uncorrected mean LipidAgeAcceleration (LAA) of 0.24, and healthy people a mean LAA of −5.33 years (Sup. Fig. 1). To make healthy people the reference when talking about LAA, we center the predictor by adding the absolute value of the mean (5.33 years) of the healthy people to each calculation. Because conversion of the underlying hazard-based signal into age-equivalent LipidAge and LAA units depends on an assumed mortality-doubling-time (MDT) parameter, we tested the stability of this scaling across a plausible MDT range. As expected, the absolute scaling of LAA and its associated hazard ratios changed modestly across different MDT values (Sup. Tab. 5), while the age-correlation structure and the relative separation between healthy, PDAC, and pancreatitis cohorts remained stable (Sup. Tab. 6).

Correspondingly, we find that PDAC patients show a mean and median lipid age acceleration of 5.57 and 4.42 years, respectively, compared to healthy people (Fig. 1B). On a population level, these differences are highly statistically significant (p= 1.46e-09). This is consistent with prior aging clock studies showing accelerated aging for cancer patients(44). Similarly, we find that the 22 patients with pancreatitis show 2.71 years mean acceleration with borderline significance compared to healthy people, and a mean deceleration of 2.86 years compared to cancer (Fig. 1B). However, due to the small sample size of the pancreatitis cohort, these findings should be considered exploratory.

### Continuous lipid age acceleration is associated with worse overall and progression-free survival

To evaluate and visualize the clinical relevance of the lipidomic clock, we stratified patients into balanced risk tiers based on baseline Lipid Age Acceleration (LAA) tertiles (Low LAA (n=48), Mid LAA (n=48), and High LAA (n=49)), while retaining continuous LAA as the primary inferential variable in the survival analyses. Kaplan-Meier survival analysis revealed a stepwise divergence in clinical outcomes across these discrete strata (Fig. 2). This association was particularly pronounced for overall survival (OS), where patients in the High LAA cohort exhibited an accelerated mortality trajectory compared to the intermediate Mid LAA and protective Low LAA cohorts, achieving a highly significant group-wise separation (multivariate log-rank p=1.23×10^−7^; Fig. 2A). A directionally consistent and robust association was similarly observed for progression-free survival (PFS), with elevated biological lipid aging correlating with significantly shortened progression-free intervals over the observation window (multivariate log-rank p=0.00139; Fig. 2B). This separation across discrete clinical tiers strongly supports the primary unadjusted continuous Cox proportional hazards modeling, where each 1-year increase in continuous LAA consistently tracks with an elevated risk of both overall mortality (HR = 1.06, p<0.001) and disease progression (HR = 1.03, p=0.019; Supplementary Fig. 2). Together, these survival metrics indicate that accelerated biological lipid aging is associated with poorer clinical outcomes.

**Figure 2:**
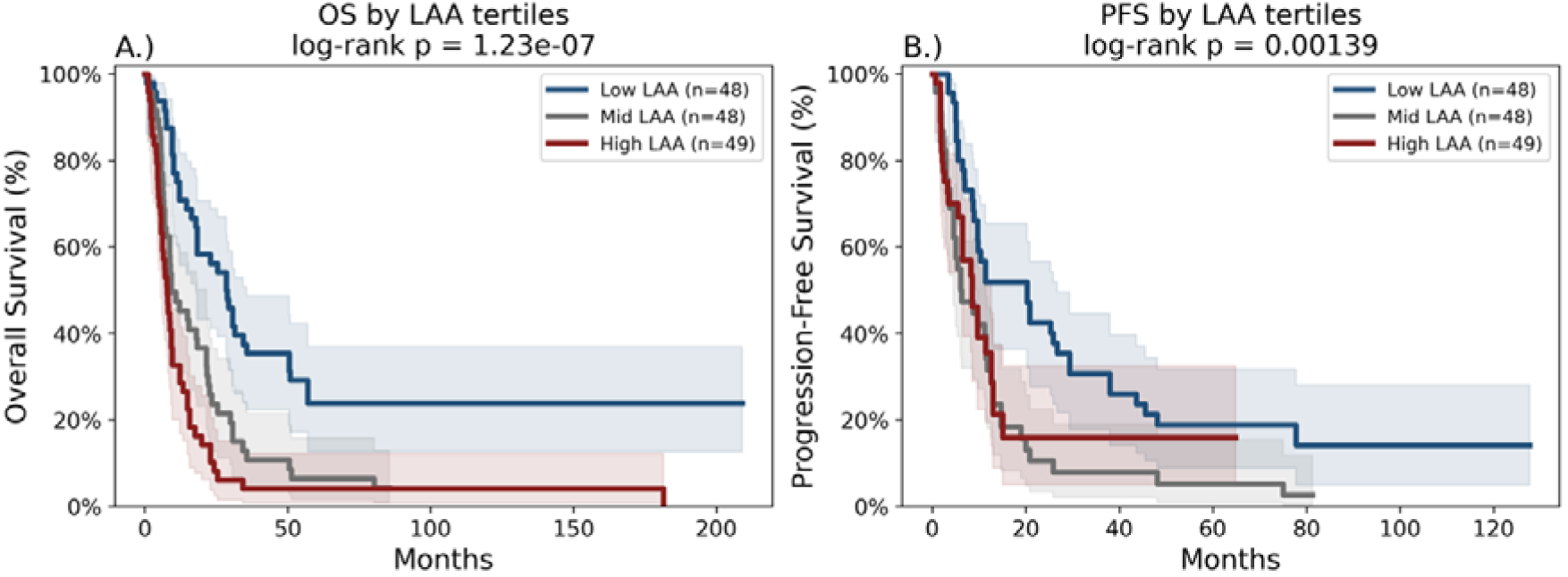
Kaplan-Meier survival analysis stratified by Lipid Age Acceleration (LAA) tertiles. Patient cohorts are stratified into three balanced risk tiers based on baseline LAA levels: Low LAA (n=48), Mid LAA (n=48), and High LAA (n=49), and primary inferential analyses were performed using continuous LAA. Shaded semitransparent bands surrounding each step function denote the corresponding 95% confidence intervals. A.) Overall Survival (OS) stratified by LAA tiers. Kaplan-Meier curves demonstrate a highly significant, stepwise divergence in overall survival clinical outcomes based on lipid biological age advancement. Patients in the High LAA cohort exhibit accelerated mortality compared to the intermediate Mid LAA and protective Low LAA cohorts (multivariate log-rank p=1.23×10^-7^). B.) Progression-Free Survival (PFS) stratified by LAA tiers. Elevated biological lipid aging consistently correlates with significantly shortened progression-free intervals, with the High LAA group showing the lowest survival probability over the 120-month observation window (multivariate log-rank p=0.00139).

### LAA remains associated with overall survival after multivariable adjustment

The adverse association of baseline Lipid Age Acceleration (LAA) with overall survival (OS) remained resilient across multiple rigorous multivariable adjustment strategies (Table 1). In the crude unadjusted univariable model across the full cohort (Model 1: N=145), LAA was significantly associated with an increased risk of death (HR = 1.063, 95% CI: 1.042–1.084, p<0.005). This effect was consistently preserved when restricting the baseline analysis to the clinical-covariate matched sub-cohort (Model 1.1: N=85, HR = 1.044, 95% CI: 1.019–1.071, p<0.005).

Following adjustment for core clinical covariates including chronological age, sex, BMI, and diabetes status, elevated LAA remained an independent risk factor (Model 2: HR = 1.093, 95% CI: 1.056–1.132, p<0.005). Crucially, adding the classical oncological biomarker CA19-9 to this clinical core did not eliminate the LAA signal, which maintained clear independent prognostic value (Model 3: HR = 1.070, 95% CI: 1.031–1.111, p<0.005) while expanding the global discriminatory power of the model (C-index = 0.74). The independent prognostic signal of LAA was further challenged by sequentially introducing heavy indicators of oncological disease burden. LAA retained strong, independent statistical significance when adjusted for distant metastasis status (Model 4: HR = 1.090, 95% CI: 1.044–1.114, p<0.005), combined metastasis and tumor grade (Model 5: HR = 1.075, 95% CI: 1.019–1.130, p<0.005), and full TNM staging criteria (Model 6: HR = 1.089, 95% CI: 1.026–1.155, p<0.005). Notably, when fitting a comprehensive complete-case model containing core clinical parameters alongside full TNMG tumor burden simultaneously (Model 7), the LAA coefficient attenuated and lost statistical significance (HR = 1.058, 95% CI: 0.983–1.137, p = 0.124), despite the global model achieving a high concordance index (C-index = 0.83). However, this model was limited by substantial clinical missingness in granular staging data, reducing the evaluable cohort from 85 to 37 patients. To assess the robustness of the association while preserving sample size, we additionally fitted a missing-indicator sensitivity model (Model 8), in which the association of LAA with overall mortality remained significant (HR = 1.072, 95% CI: 1.030–1.115, p < 0.005). These findings support the overall robustness of the LAA signal, but they do not fully resolve the limitation imposed by missing granular staging data in the complete-case TNMG model.

To further rule out systemic clinical confounding, we performed a treatment-status sensitivity analysis. Baseline LAA levels did not differ significantly between patients who had received prior therapy and those in the pretreatment group (Sup. Fig. 3). Furthermore, explicitly adjusting for treatment status in the final survival models did not alter or remove the independent LAA survival association (HR=1.09, 95% CI: 1.05-1.13, p<0.005; Sup. Tab. 7), confirming that the prognostic value of the clock remained significant despite variations in therapeutic timing. Similar independent trends for LAA were observed across most progression-free survival models, even though generally the signal was weaker (Sup. Tab. 8).

**Figure 3:**
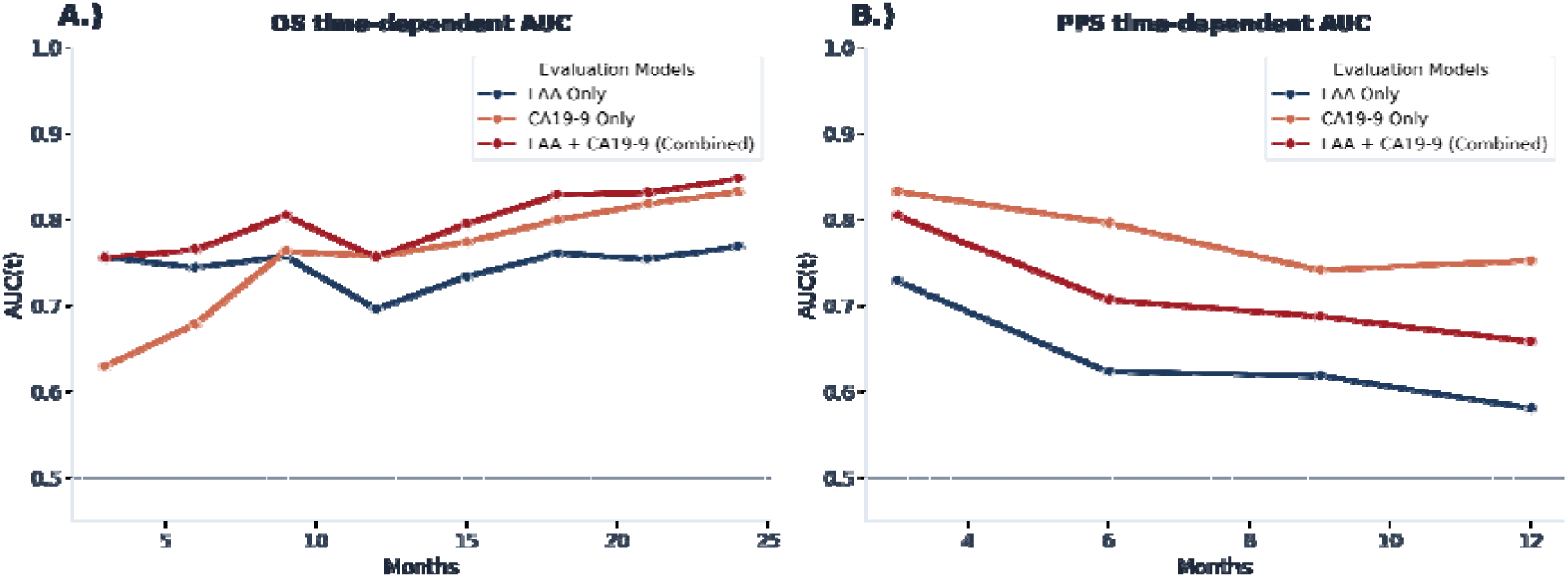
Time-dependent area under the receiver operating characteristic curve (AUC) analysis evaluating the prognostic accuracy of Lipid Age Acceleration (LAA) and CA19-9. Longitudinal prognostic performance is shown for three distinct evaluation frameworks: LAA model alone, CA19-9 biomarker alone, and the integrated model combining LAA and CA19-9. The dashed horizontal line at 0.5 denotes random classification performance. A.) Longitudinal performance for Overall Survival (OS). Predictive accuracy evaluated over a 24-month clinical follow-up period. The combined model (LAA + CA19-9) consistently demonstrates superior discriminative performance across nearly all time points, achieving a peak longitudinal AUC of 0.85 at 24 months and a superior mean longitudinal accuracy (mean AUC = 0.80) relative to CA19-9 alone (mean AUC = 0.76) and LAA alone (mean AUC = 0.75). B.) Longitudinal performance for Progression-Free Survival (PFS). Discriminative capacity tracked over a 12-month interval. For PFS prediction, CA19-9 alone displays the highest baseline and longitudinal capacity (mean AUC = 0.78), followed by the integrated model (mean AUC = 0.71), while LAA alone maintains modest but stable predictive performance above chance (mean AUC = 0.64).

### Time-dependent AUC analysis supports complementary OS discrimination

To evaluate the longitudinal profile of predictive performance, a time-dependent area under the receiver operating characteristic curve [AUC(t)] analysis was conducted across sequential follow-up horizons (Fig. 3). For overall survival (OS), discriminative accuracy remained consistently well above random chance (AUC = 0.50) across all evaluated time points for both individual and combined feature sets. Standing alone, Lipid Age Acceleration (LAA) demonstrated meaningful independent OS discrimination, achieving a mean AUC of 0.747 and a median AUC of 0.756 across the follow-up window. Crucially, the combined framework (LAA + CA19-9) yielded the strongest overall discrimination across the majority of the clinical follow-up window, demonstrating a mean longitudinal AUC of 0.798 and a median AUC of 0.800. This additive effect was particularly pronounced at intermediate and extended time horizons, where the combined model consistently outperformed the individual metrics, ultimately reaching a peak longitudinal AUC of 0.85 at the 24-month threshold. This additive pattern was not observed for progression-free survival. For PFS, CA19-9 showed stronger discrimination than LAA. (mean AUC = 0.781). Standalone LAA performance was markedly weaker (mean AUC = 0.638), and its integration into the joint model failed to provide an additive benefit, instead attenuating the standalone predictive power of CA19-9 to a combined mean AUC of 0.715. These time-dependent metrics demonstrate that the predictive utility of the lipidomic clock is highly phenotypic and specific to overall mortality dynamics rather than reflecting a broad, non-specific signature across all oncology endpoints.

### Bootstrap ridge stability highlights a reproducible sphingolipid-associated component of the clock

Because the predictive lipid model favored Ridge regularization—which shrinks coefficients uniformly rather than enforcing mathematical sparsity—traditional feature selection frequencies do not fully capture model robustness. Instead, feature stability was comprehensively evaluated by tracking the Top-30 recurrence frequency of individual lipids, their directional sign consistency, and class-level aggregation across 500 independent bootstrap refits (Fig. 4). This approach demonstrated that the predictive signal of the clock is highly structured; rather than reflecting stochastic noise, a distinct subset of lipids consistently emerged as primary drivers of the clock, with class-level recurrence anchoring heavily around sphingolipids—specifically sphingomyelins (SM) and ceramides (Cer)(Sup. Fig. 4). When evaluating the individual lipid contributors in the final multivariable model, this structural robustness was confirmed, as exactly two species achieved independent frequentist statistical significance: SM 41:2 (p = 0.022) and TG 58:2 (p = 0.027) (Sup. Tab. 9). Importantly, the significance of these two lipids in the final model converges with the bootstrap stability metrics, validating them as the anchors of the lipidomic clock. Specifically, SM 41:2 emerged as the single most recurrent feature across all 500 bootstrap resamples (Fig. 4A) and demonstrated absolute directional reliability (sign consistency = 1.00) toward a negative coefficient (Fig. 4B), mirroring its significant protective association against biological age acceleration in the final model. TG 58:2 similarly exhibited absolute directional reliability (sign consistency = 1.00) across bootstrap iterations, consistently maintaining a positive coefficient that directly aligns with its independent risk profile and formal statistical significance in the final model as an indicator of accelerated lipid aging. While other lipid species (such as PC 36:2 and SM 34:2) displayed high directional uniformity during the bootstrap resampling phase, they did not reach significance when fully adjusted in the final model. This tight convergence between our bootstrap stability selection and the final model highlights a reproducible lipidomic signature driving the aging clock, rather than an artifact of cohort-specific noise.

**Figure 4:**
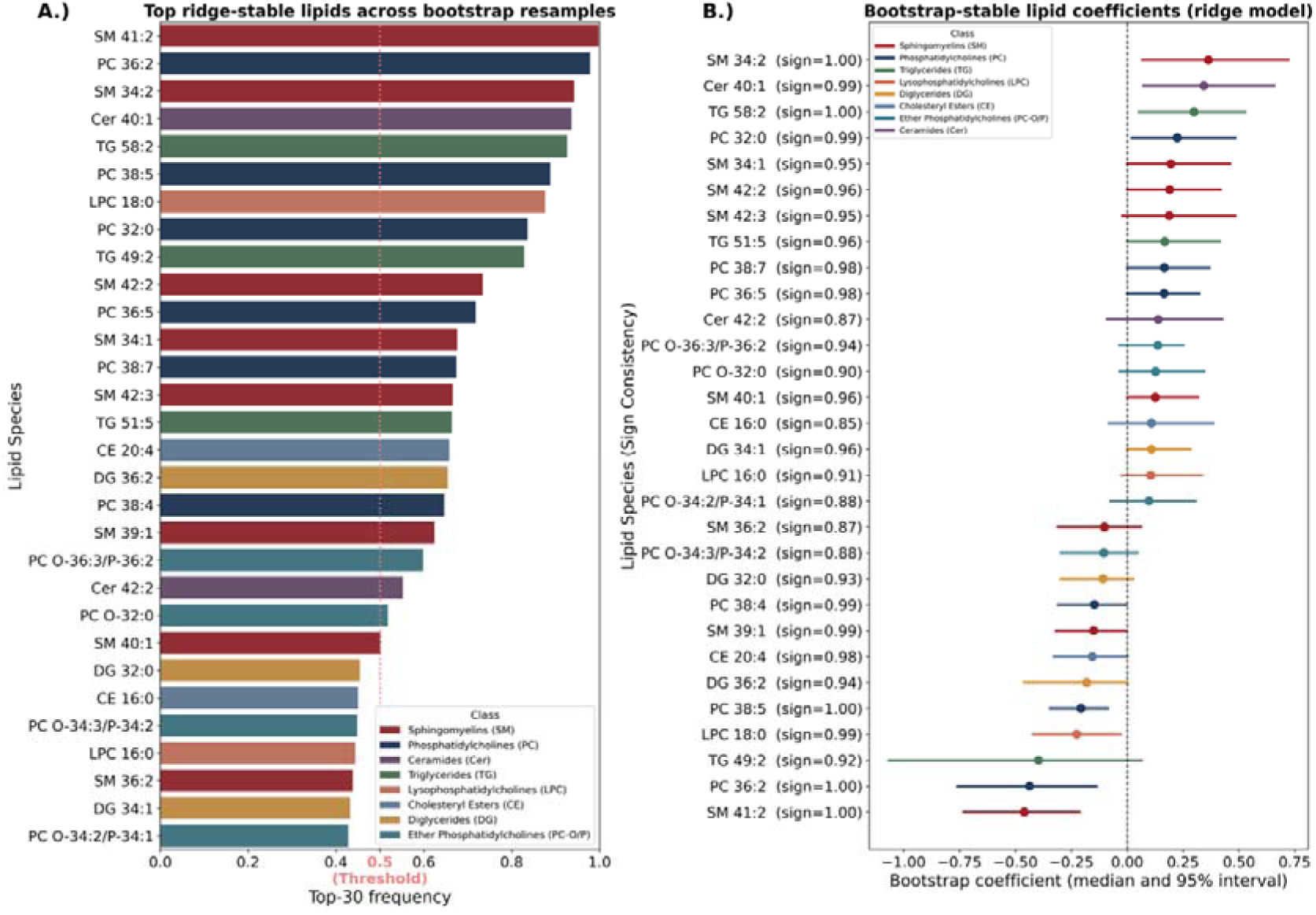
Bootstrap stability selection and coefficient robustness of the lipidomics-based aging clock. This figure details the feature selection architecture and parameter stability using iterative bootstrap resampling within the penalized ridge regression framework. A.) Selection frequency of top ridge-stable lipid species. The horizontal bar plot illustrates the proportion of bootstrap resamples (0.0 to 1.0) in which a given lipid species was selected among the top 30 most predictive features. Bars are color-coded by major lipid classes according to the structural legend. The vertical dashed red line denotes the inclusion threshold of 0.50, demonstrating that the prioritized biomarkers exhibit highly reproducible predictive power independent of sample shuffling. B.) Distribution and directional stability of bootstrap-stable lipid coefficients. The forest plot displays the median effect size and corresponding 95% confidence intervals (intervals) for the ridge regression coefficients across all bootstrap iterations. The vertical dashed black line at 0.0 separates positive coefficients (right; indicating lipids positively associated with accelerated biological aging) from negative coefficients (left; indicating lipids inversely associated with age acceleration). Numerical values alongside the y-axis labels denote sign consistency (e.g., sign = 1.00), confirming the high directional uniformity of the selected feature weights across the resampled ensembles.

### A reduced clock based on complex lipids captures the aging signal with fewer features

To investigate whether the predictive power of the lipidomic clock is distributed across the entire lipidome or concentrated within specific functional domains, we constructed a streamlined, parsimonious model restricted exclusively to complex structural and signaling lipids, such as phospholipids and sphingolipids, while completely omitting neutral storage lipids like triacylglycerols and diacylglycerols. This focused Complex LipidClock framework utilized fewer than half the features of the original unconstrained architecture, shrinking the biomarker panel by approximately 46% from 102 down to 55 features, yet it successfully captured the essential biological aging and prognostic signals (Tab. 2). In head-to-head survival comparisons, the parsimonious complex clock achieved independent prognostic performance that was remarkably robust and virtually identical to the full model. Specifically, the Complex LipidClock maintained an equivalent effect size for progression-free survival (HR = 1.028, p=0.032) and demonstrated a marginally stronger hazard ratio for overall survival (HR = 1.067, p=5.65×10^−9^) compared to the full 102-feature model (OS HR = 1.063, p=1.72×10 ^−9^). Beyond preserving robust survival stratification, the streamlined complex clock exhibited enhanced sensitivity when differentiating benign inflammatory conditions from healthy baselines (Sup. Fig. 5). While the full clock model only captured a marginal, non-significant trend toward lipid age acceleration in pancreatitis patients (p=0.0653), the 55-feature complex lipid framework revealed formal, statistically significant biological age acceleration within this cohort relative to healthy controls (p=0.0482). Meanwhile, the highly pronounced age acceleration signal characteristic of the PDAC cohort remained distinct (p=1.14×10^−9^). Taken together, these findings indicate that much of the aging-relevant and prognostic signal in this dataset appears recoverable from structural and signaling complex lipids, rather than neutral lipids. These findings support the feasibility of future targeted biomarker assays focused on the complex lipidome to achieve greater clinical parsimony without sacrificing diagnostic or prognostic precision.

## Discussion

In this study, we developed and internally evaluated the first oncological biological aging clock derived from the serum lipidomic profiles of 443 human subjects with pancreatic ductal adenocarcinoma. Despite having been trained exclusively on the lipidome and mortality data of an active malignant cohort, the clock demonstrated high accuracy when deployed on healthy individuals and pancreatitis samples from the same parent dataset (Fig. 1). A key conceptual finding of this framework is that the lipid age predictor systematically assigned a lower biological age to healthy volunteers. This divergence highlights an overlap between lipid signatures associated with mortality in PDAC and lipidomic patterns previously linked to natural human aging.

Our findings indicate that aging and mortality signatures remain highly conserved across the spectrum from severe, life-threatening malignancies to non-diseased baseline states. In this sense, clinical oncology cohorts could be effectively repurposed to develop systemic aging biomarkers. This paradigm is particularly valuable for translational research because oncology and chronic disease biobanks routinely contain substantially larger sample numbers and more complete longitudinal survival outcomes than datasets composed purely of healthy agers.

The baseline synchronization of these dynamics is further supported by our correlation analyses. The Pearson correlation coefficient between chronological age and predicted LipidAge was 0.86 (p=2.92×10^−78^) for healthy controls and 0.72 (p=3.17×10^−24^) for PDAC patients (Fig. 1). This indicates that systemic aging and cancer-driven metabolic decay are heavily correlated, suggesting that metabolic processes which normally unfold across several decades during natural aging are severely compressed in cancer into a much shorter, accelerated clinical timescale.

Because PDAC is an exceptionally aggressive disease with a 5-year survival rate typically remaining under 10%(43), chronological age itself is not strongly predictive of survival within the cohort; the malignancy acts as the dominant driver of mortality almost independently of a patient’s time alive. This aligns with clinical literature reporting that short- and intermediate-term survival outcomes for PDAC remain highly similar across disparate age groups, confirming that chronological age is a poor determinant of individual host resilience(51). In contrast, our LipidAge-based metrics demonstrate robust predictive power for PDAC survival endpoints. When evaluated as a continuous variable, each 1-year increase in Lipid Age Acceleration (LAA) significantly escalates the hazard of death, and this prognostic signal remains independent of traditional oncological risk parameters (Tab. 1). Hence continuous LAA remained a significant predictor of overall mortality even after adjusting the model for standard clinical cancer measures, staging parameters, and the conventional tumor marker CA19-9. This preservation of effect size indicates that the lipid clock captures mortality-associated lipidomic variation not fully reflected by traditional tumor-centric staging metrics and may partly reflect broader systemic features related to host state or physiological resilience. To visually and clinically illustrate this risk distribution, we stratified the cohort into discrete LAA risk tertiles (Low, Mid, and High LAA). Kaplan-Meier survival curves revealed a highly significant stepwise separation in clinical trajectories (Fig. 2). This graded divergence was evident for overall survival (multivariate log-rank p=1.23×10^−7^), with the High LAA group exhibiting the sharpest, most rapid decline in survival probability, alongside a directionally consistent separation for progression-free survival (multivariate log-rank p=0.00139). This robust stratification validates our continuous multivariable modeling and confirms that accelerated biological lipid aging tracks tightly with unfavorable clinical endpoints. Our 500-fold bootstrap stability selection framework converged directly with our final multivariable regression to identify a reproducible lipid signature driving the clock. Two specific lipid species that achieved statistical significance: SM 41:2 (p=0.022) and TG 58:2 (p=0.027). Well-regulated sphingolipids are established structural and signaling hallmarks of overall health and longevity, whereas their systemic dysregulation has been universally linked to metabolic disease, chronic inflammation, and accelerated mortality(52–54). In our analysis, SM 41:2 emerged as a powerful protective anchor, displaying absolute directional reliability across all 500 bootstrap iterations, which directly mirrors its significant association with extended survival in the final model. Conversely, the neutral storage lipid TG 58:2 consistently maintained a positive coefficient, confirming its role as a stable indicator of advanced biological aging and elevated risk. Prior longevity and epidemiological studies have consistently linked these identical lipid classes to centenarian cohorts, familial longevity, physical function, frailty, and multi-morbidity metrics, further anchoring our clock’s selections within established geroscience paradigms(15,16,53,55–61). The structural importance of sphingomyelins and ceramides within the clock architecture points directly toward a mechanistic connection with cellular aging and systemic inflammation. Other geroscience investigations have highlighted the critical role of sphingolipids in human aging trajectories, for instance, in the Baltimore Longitudinal Study of Aging and other cohorts, plasma sphingomyelins were shown to undergo significant, age-dependent alterations(62–64).

Impaired sphingolipid metabolism is involved in a lot of pathophysiological phenotypes(11). Sphingolipids, especially ceramides and sphingosine-1-phosphate, regulate cellular processes crucial for immune responses, inflammation(65), as well as cell growth and survival(66).

Moreover, ceramide species undergo strong changes with age(67) and have been proposed as biomarkers of human longevity(68).

PDAC cell lines maintain altered sphingolipid signaling and elevated sphingosine-1-phosphate to preserve cancerous phenotypes(69). In addition, cellular senescence, a hallmark of aging(70), can be induced by ceramides, whereas sphingosine-1–phosphate can delay the transition to a senescent state(71). More broadly, recent work has emphasized that cellular aging and senescence are central determinants of cancer cell fate and tumor-associated biological heterogeneity(72), further supporting the relevance of senescence-linked lipid pathways in PDAC.

Finally, sphingolipids are deeply involved in inflammation and disease progression(52,54,65). As our LipidClock shows enrichment in sphingolipids, we argue that a substantial component of the LipidClock may reflect lipidomic patterns consistent with inflammatory and immune-related processes linked to aging and disease(73), although these biological processes were not directly measured by the model. This interpretation is in line with other aging clocks that highlight immune dysfunction and inflammation as central features of aging, including glycan-based clocks and iAge(24,36,74).

In our case, this inflammatory sensitivity was clearly demonstrated when we streamlined our framework into a parsimonious 55-feature Complex LipidClock. By omitting neutral storage lipids entirely and focusing exclusively on structural and signaling complex pathways, we reduced the variable burden by 46% with similar performance in prognostic survival accuracy. Remarkably, this refined clock showed statistically significant acceleration in this exploratory pancreatitis cohort relative to healthy controls (p=0.0482), a distinction that was obscured as a non-significant trend in the unconstrained 102-feature model (p=0.0653). Overall, the LipidClock provides evidence that complex lipid metabolism is deeply intertwined with survival and mortality, both in natural human aging and in aggressive diseases of accelerated biological aging such as PDAC. This tight convergence allows the accurate deployment of circulating lipidomic signatures to track biological age, demonstrating that the lipidomic dynamics of natural aging mirror the metabolic decay observed in cancer close to death, where homeostatic failure is accelerated and processes that normally require decades to break down are compressed into years.

As large scale lipidomic data with thousands of individual samples is still hard to come by a major limitation of our study is that we build our model on only 443 human serum samples. While this is enough to gain insights into some dynamics of cancer and aging, it is not enough to capture the entire complexity of these biological processes. Although the model generalized across held-out PDAC samples and across healthy and pancreatitis groups within the same study, all analyses were performed within a single parent dataset and analytical platform. Independent external cohorts will therefore be needed to determine the broader transportability and generalizability of the present findings.

Nevertheless, what this study shows is that lipids open an avenue for cancer and aging biomarkers using aging clock methodology. However, at present, LAA should be interpreted as a candidate prognostic or risk-stratification marker rather than as a guide for treatment selection. Although higher LAA was associated with poorer survival and retained predictive value after multivariable adjustment, the present study did not evaluate treatment toxicity, therapeutic response, calibration, net benefit, or prospective clinical utility. These questions will require dedicated validation in future cohorts.

Future work should focus on larger lipidomic datasets, broader lipid coverage, and lipid-class-specific clocks, with sphingolipids being a prime contender. Lipid aging clocks could be used in future oncological or longevity clinical trials to assess the safety and efficacy of therapeutic drugs against pancreatic ductal adenocarcinoma, other cancers, and longevity(75,76).

### Conclusion

This study describes a novel pathology-trained lipid aging clock that successfully links pancreatic disease to systemic, inflammation-associated biological age acceleration. We demonstrate that advanced baseline Lipid Age Acceleration acts as a predictor associated with shortened overall and progression-free survival in pancreatic ductal adenocarcinoma, capturing critical prognostic information complementary to traditional tumor markers like CA19-9. Furthermore, our analysis supports the interpretation that biological aging and cancer-associated mortality may share overlapping lipidomic features, particularly involving sphingolipid and complex-lipid pathways. However, because the model was developed and evaluated within a single parent dataset, independent external validation will be required before broader generalizability can be concluded.

Exploratory analyses of the small pancreatitis sub-cohort suggested similar lipid age acceleration, but these findings require confirmation in larger independent cohorts. Ultimately, this work supports the feasibility of deriving aging-relevant lipidomic clocks from pathological datasets, providing a clear path forward for targeted, clinically parsimonious lipidomic assays capable of monitoring human aging, systemic inflammation, and disease-associated mortality.

## Data Availability Statement

All the data is uploaded on GitHub at https://github.com/max-unfried/pdac-lipidclock/

## Code Availability Statement

All the code is available at github at https://github.com/max-unfried/pdac-lipidclock/

## Author contributions

Maximilian Unfried: Conceptualization, methodology, data analysis, and writing – original draft preparation.

Amaury Cazenave-Gassiot: Data Collection, and writing – review and editing. Evelyne Bischof: Writing – review and editing.

Michal Holcapek: Data Collection, and writing – review and editing.

Morten Scheibye-Knudsen: Supervision, and writing – review and editing.

Markus R. Wenk: Conceptualization, supervision, and writing – review and editing. Jan Gruber: Conceptualization, supervision, methodology and writing – review and editing.

Brian K. Kennedy: Conceptualization, supervision, and writing – review and editing

