## Supplementary Info for "A Pathology-Trained Lipid Aging Clock Reveals Accelerated Aging and Prognostic Sphingolipid Signatures in Pancreatic Disease"

This file provides additional information on data preprocessing, lipid quantification methods, statistical modeling, and supplementary analyses not included in the main manuscript.

**Supplementary for Material and Methods**

**Demographics of the Dataset**

In total, 830 samples were available in the dataset of which 262 were healthy, 546 were diagnosed with pancreatic ductal adenocarcinoma, and 22 with pancreatitis. Of the PDAC patients 443 had a recorded overall – and 439 progression free survival. The mean age for healthy people was 53.02 +/- 11.88 years with a range of 19-79 years, for PDAC patients it is 63.85 +/- 10.2 on a range from 23-87 years, and 61.27+/-11.89 with a range of 37-75 years for people with pancreatitis. Age distribution between healthy and PDAC, and healthy and pancreatitis is statistically significantly different (Welch’s t-test: p<0.005), but not between PDAC and pancreatitis groups (Welch’s t-test: p>0.05 for both). The distribution of sex did not differ significantly across the healthy controls, pancreatitis, and PDAC cohorts (χ^2^ (2,N=830)=0.89,p=0.641). (Sup. Tab. 1).

|  | **Healthy** | | **PDAC** | | **Pancreatitis** | |
| --- | --- | --- | --- | --- | --- | --- |
| **Total Count** | 262 | | 546 | | 22 | |
| **Overall Survival Information Count** | N/A | | 443 | | N/A | |
| **Progression Free Survival Information Count** | N/A | | 439 | | N/A | |
| **OS and PFS count** | N/A | | 439 | | N/A | |
| **CA19-9 Count** | 262 | | 544 | | 22 | |
| **Diabetes Count** | 34 | | 210 | | 7 | |
| **Mean Age in years** | 53.02 +/- 11.88 | | 63.85 +/- 10.2 | | 61.27+/-11.89 | |
| **Age Range in years** | 19-79 | | 23-87 | | 37-75 | |
| **Sex** | Male | Female | Male | Female | Male | Female |
| **Count per Sex** | 128 | 134 | 275 | 271 | 13 | 9 |

*Sup Tab. 1: Dataset Demographics*

**Collected Lipid Species**

Detailed information on lipid extraction and processing can be found in Wolrab *et al.*^1^ Briefly, in Phase III (verification) of said study, a total of 202 individual lipid species from 11 classes (Sup. Tab. 3) were quantified using a UHPSFC/MS method validated in accordance with FDA and EMA guidelines^2^. Solvent blanks and pooled serum QC samples (including NIST SRM reference plasma^3^) were extracted using a modified Folch protocol including multiple class-specific internal standards^1^. Pooled QC samples were measured every 40 samples, and a mixture of naturally occurring lipid species served as a system suitability standard. Instrumental stability and sample preparation quality were monitored throughout each sequence by tracking signal responses of selected endogenous lipids and internal standards across all samples, enabling detection of outliers from preparation errors or instrumental drift. PCA of all lipidomic profiles was additionally performed to identify outliers and confirm QC sample clustering.

Candidate lipid variables were additionally screened for excessive value pile-up. Lipids were excluded if >5% of samples were at the minimum observed value or if >5% of samples shared the same exact value. Remaining lipids were then required to have complete and strictly positive values across samples, leaving us with 102 species from the original 202 species, before preprocessing and model fitting. The exclusion criterion was quite strict such that only species without missing values were used.

| **Lipid Class** | **Measured lipids** | **Lipids used for analysis in full clock** | **Lipids used for analysis in complex lipid clock** |
| --- | --- | --- | --- |
| Triglycerides (TG) | 84 | 42 | 0 |
| Phosphatidylcholines (PC) | 25 | 17 | 17 |
| Sphingomyelins (SM) | 23 | 13 | 13 |
| Diglycerides (DG) | 22 | 5 | 0 |
| Ether Phosphatidylcholines (PC-O/P) | 15 | 9 | 9 |
| Ceramides (Cer) | 11 | 4 | 4 |
| Cholesteryl Esters (CE) | 9 | 7 | 7 |
| Lysophosphatidylcholines (LPC) | 8 | 5 | 5 |
| Monoglycerides (MG) | 3 | 0 | 0 |
| Cholesterol (Chol) | 1 | 0 | 0 |
| Coenzyme Q10 | 1 | 0 | 0 |
| Total | 202 | 102 | 55 |

*Sup Tab. 2:* *Lipid Classes Count for the various lipid classes*

**Data Preprocessing**

Candidate lipid variables were defined after removal of non-lipid metadata columns. For primary clock construction, only lipid species with valid positive numeric values in the relevant analytical datasets were retained, missing lipid values were not imputed, resulting in 102 lipids to be used (Sup. Tab. 2). For each retained lipid, the median abundance was calculated in the PDAC training cohort only. Sample-level abundances in the training set, held-out PDAC test set, healthy controls, and pancreatitis samples were then normalized to these training-set medians, and the resulting fold-change values were log2-transformed prior to modelling. This training-only normalization strategy was used to reduce skewness, improve comparability across lipids, and avoid information leakage from non-training samples into feature scaling.

**Dataset Partitioning and Stratification and Demographics of the train-test split**

To ensure a balanced and robust framework for model training and evaluation, the primary analysis cohort containing concurrent overall survival (OS) and progression-free survival (PFS) data (n=439) was partitioned into independent training and testing sets using a stratified split configuration (67% training, 33% testing) (Sup. Tab. 3). Stratification was strictly enforced across two critical clinical dimensions: patient age and overall survival outcomes. To capture non-linear age effects, continuous age values were discretized into four equal-sized bins (quartiles) prior to partitioning, ensuring uniform chronological representation. Concurrently, stratification by survival event status was integrated to maintain an identical event-to-censor ratio and preserve the underlying survival curves across both subsets. The demographics of the train-test split can be found in Sup. Tab. 3. Following the split, data balance was rigorously verified using post-hoc statistical testing. Pairwise comparisons confirmed no statistically significant differences between the training and testing cohorts with respect to age distribution (Welch's t-test, p>0.05), sex distribution (χ^2^ test of independence, p>0.05), or baseline diabetes prevalence (χ^2^ test, p>0.05). This statistical alignment confirms an unbiased allocation of clinical characteristics, minimizing confounding risks during model evaluation.

|  | **Train (67%)** | | **Test (33%)** | |
| --- | --- | --- | --- | --- |
| **Total Count** | 294 | | 145 | |
| **Overall Survival Information Count** | 294 | | 145 | |
| **Progression Free Survival Information Count** | 294 | | 145 | |
| **OS and PFS count** | 294 | | 145 | |
| **CA19-9 Count** | 293 | | 145 | |
| **Diabetes Count** | 102 | | 49 | |
| **Mean Age in years** | 63.43 +/- 10.21 | | 62.94 +/- 10.64 | |
| **Age Range in years** | 32–87 | | 23–81 | |
| **Sex** | Male | Female | Male | Female |
| **Count per Sex** | 152 | 142 | 70 | 75 |

*Sup Tab. 3: Demographics of the train and test set*

**Constructing the Lipid Clock**

The construction of the Lipid Clock follows *Fong et al*^4^. First, a cox proportional hazard model with chronological age as the only covariate was trained. This chronological age model provides the hazard based on chronological age ($H_{CA}$). Next, a penalized cox proportional hazard model with all the lipid species as covariates. This lipid model gives us the hazard based on the lipidome ($H_{lipid}$).

The intuition behind developing the clock like this is as follows:

By dividing the predicted hazards $H_{lipid}$ and $H_{CA}$ we get the individual ratio R of partial hazards for each individual.

$$R=\frac{H_{lipid}}{H_{CA}}$$

A ratio of 1 means that the hazard of the lipidome corresponds to the hazard of the chronological age, and thereby the lipidome is typical for the chronological age. A ratio R>1 means that the lipidome is biologically older than the chronological age, whereas a ratio R<1 would indicate a lipidome younger than the chronological age. Through a log2 transform of the ratio we introduce acceleration and deceleration, and to scale this ratio into years we multiply it by 8 - the mortality rate doubling time for humans, as described by Gompertz law of mortality^5^, which gives us a measure for Lipid Age Acceleration (LAA) defined as

$$LAA=8 years*log2 \frac{H_{lipid}}{H_{CA}}$$

With R =1, LAA is 0, for R>1 LAA leads to acceleration and LAA is decelerated for R<0.

Therein, LipidAge can then be interfered by LipidAge = Chronological Age + LAA.

As the model is trained on severely diseased individuals, it will calculate an LAA for healthy individuals that is negative. Therefore, to make LAA referenced to healthy people we use an additive transformation and mean center the LAA by adding the absolute value of the mean of the LAA of healthy people.

Hence, centered LAA is given by LAA + mean(${LAA}_{healthy}$).

**Model training and hyperparameter optimization**

We optimize the parameters for the penalized cox proportional hazard model through grid search and 5-fold stratified cross validation. Hyperparameters were tuned by stratified cross-validation within the PDAC training cohort. Stratification was performed using OS event occurrence together with age-group information to maintain comparable event and age distributions across folds, and mean concordance index was used to select the final model.

The final lipid model was ridge-like regression with penalizer=0.1, and l1-ratio of 0.05 achieved the highest cross-validated concordance index (0.68).

Grid search over Elastic Net hyperparameters for the univariate chronological age model showed near-identical cross-validated concordance indices across all tested settings (mean C-index range: 0.55). This reflects the low risk of overfitting with a single predictor. We selected penalizer=0.0001 and l1_ratio=0.0 (mean C-index=0.5537), for the age model.

**Implementation**

For the implementation Python v3 was used. Survival analysis was conducted using the Lifelines package^6^. Data, code, and model will be made available on GitHub <https://github.com/max-unfried/pdac-lipidclock>.

**Supplementary Results**

**Figures**

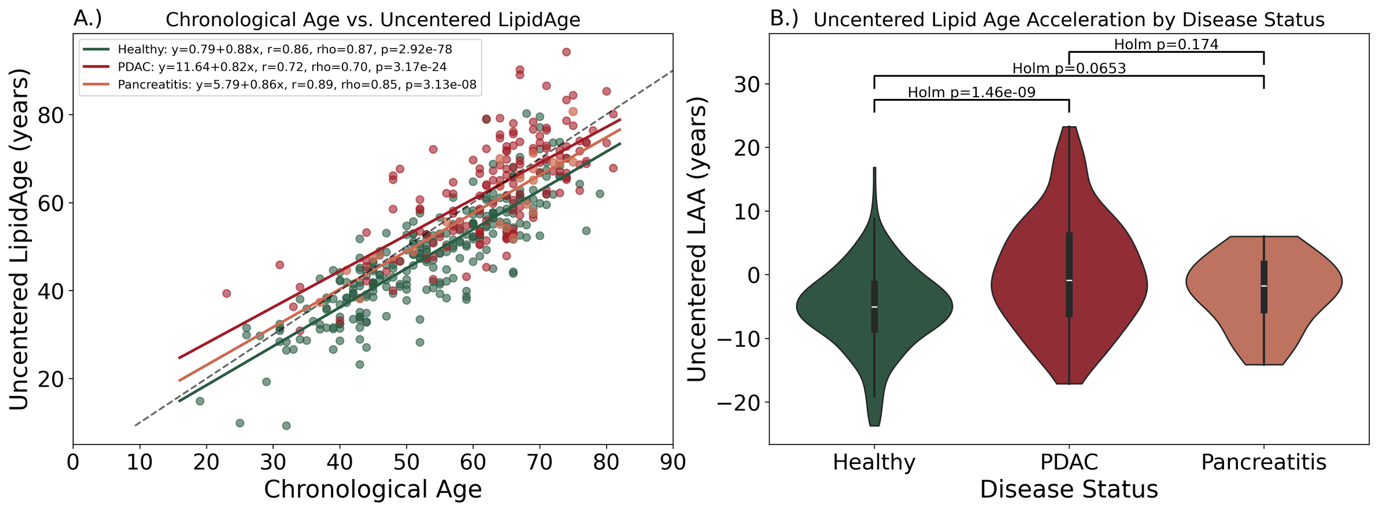

*Sup. Fig 1:* *Uncentered results for A.) Chronological age vs LipidAge for Healthy, PDAC and Pancreatitis Patients; B.) Violin plot of Lipid Age Acceleration for Healthy, PDAC and Pancreatitis Patients.*

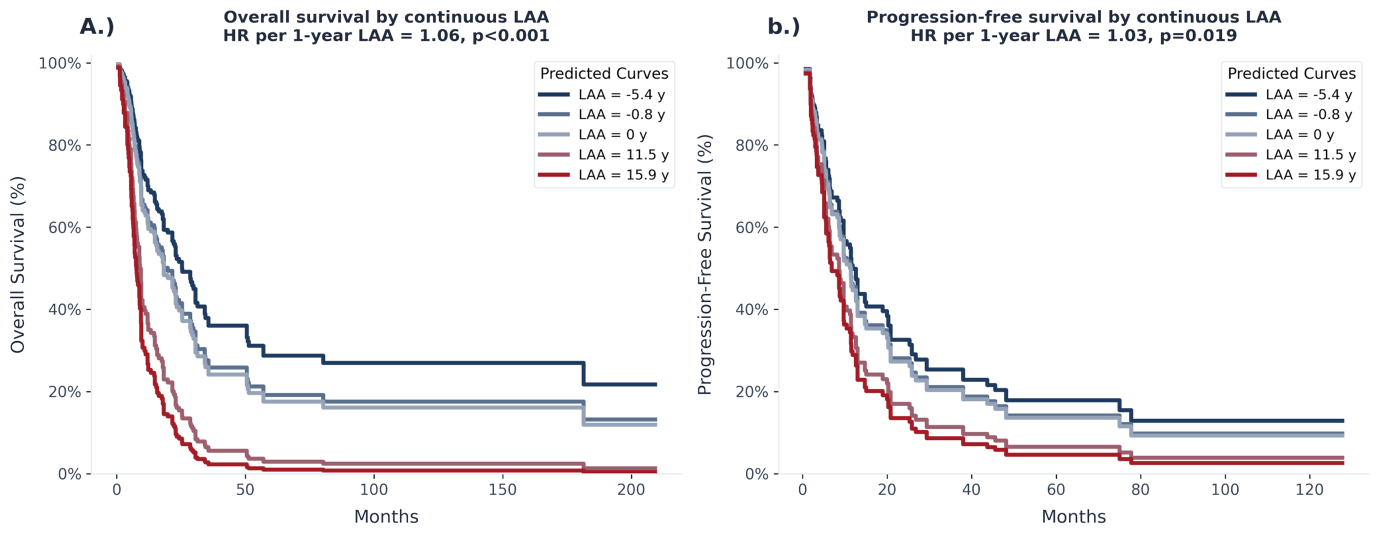

*Sup. Fig 2:. Univariable Cox model predicted survival curves by continuous LAA. Predicted survival curves illustrating the unadjusted effects of continuous Lipid Age Acceleration (LAA, measured in years) on clinical outcomes. A.) Predicted curves for overall survival (OS) by continuous LAA. Each 1-year increase in baseline LAA is associated with a significantly higher risk of overall mortality (Hazard Ratio [HR] = 1.06 per 1-year LAA, p<0.001). B.) Predicted curves for progression-free survival (PFS) by continuous LAA. Elevated baseline LAA values significantly correlate with an increased risk of disease progression or death (HR = 1.03 per 1-year LAA, p=0.019). Curves represent the predicted survival probabilities generated from univariable Cox proportional hazards models at specific continuous LAA values (–5.4 y, –0.8 y, 0 y, 11.5 y, and 15.9 y).*

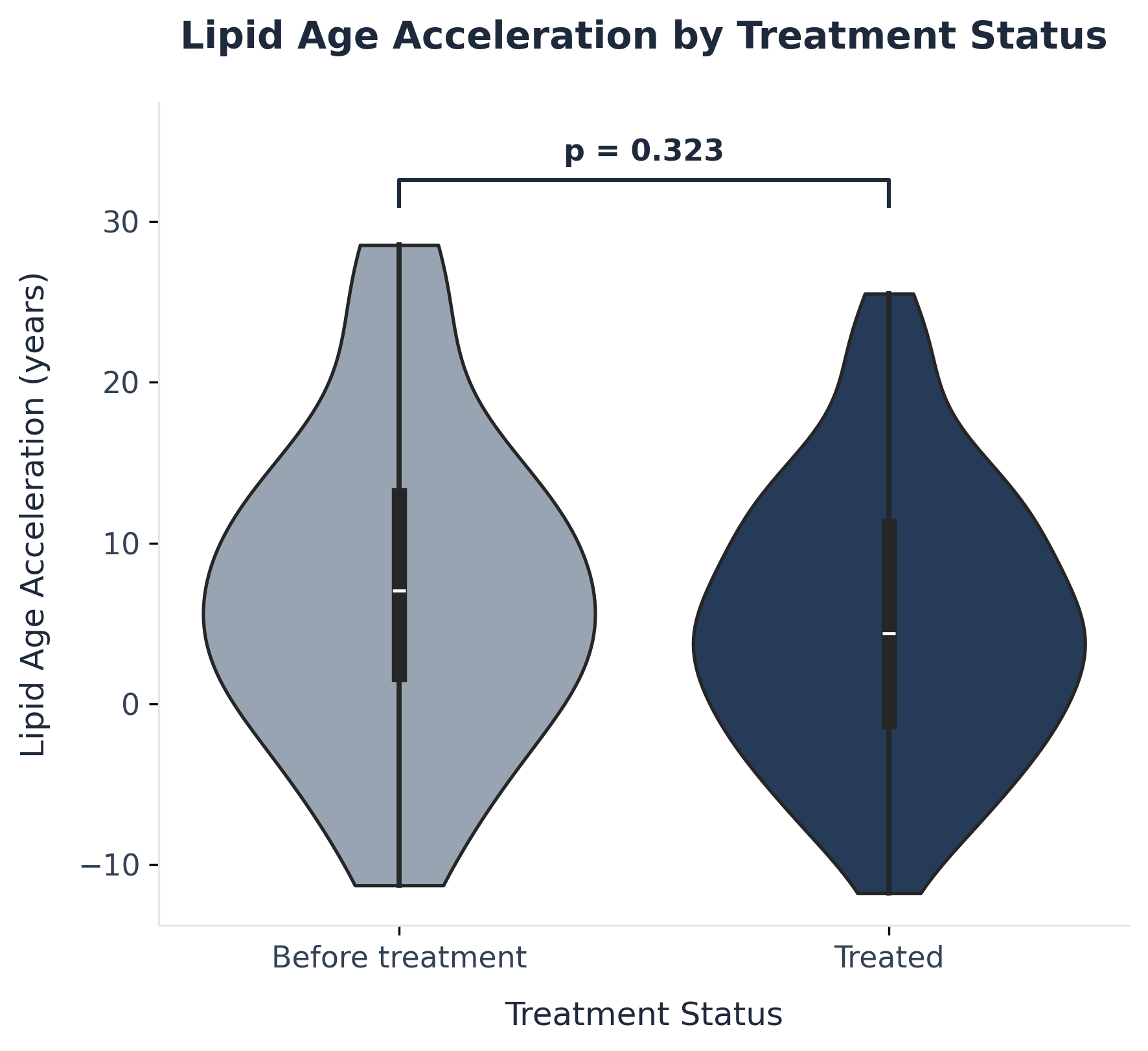

*Sup Fig. 3:* *Lipid Age Acceleration (LAA) compared by treatment status. Violin plots demonstrate no statistically significant difference in biological age acceleration between patients before treatment and after treatment (Mann-Whitney U p=0.323*

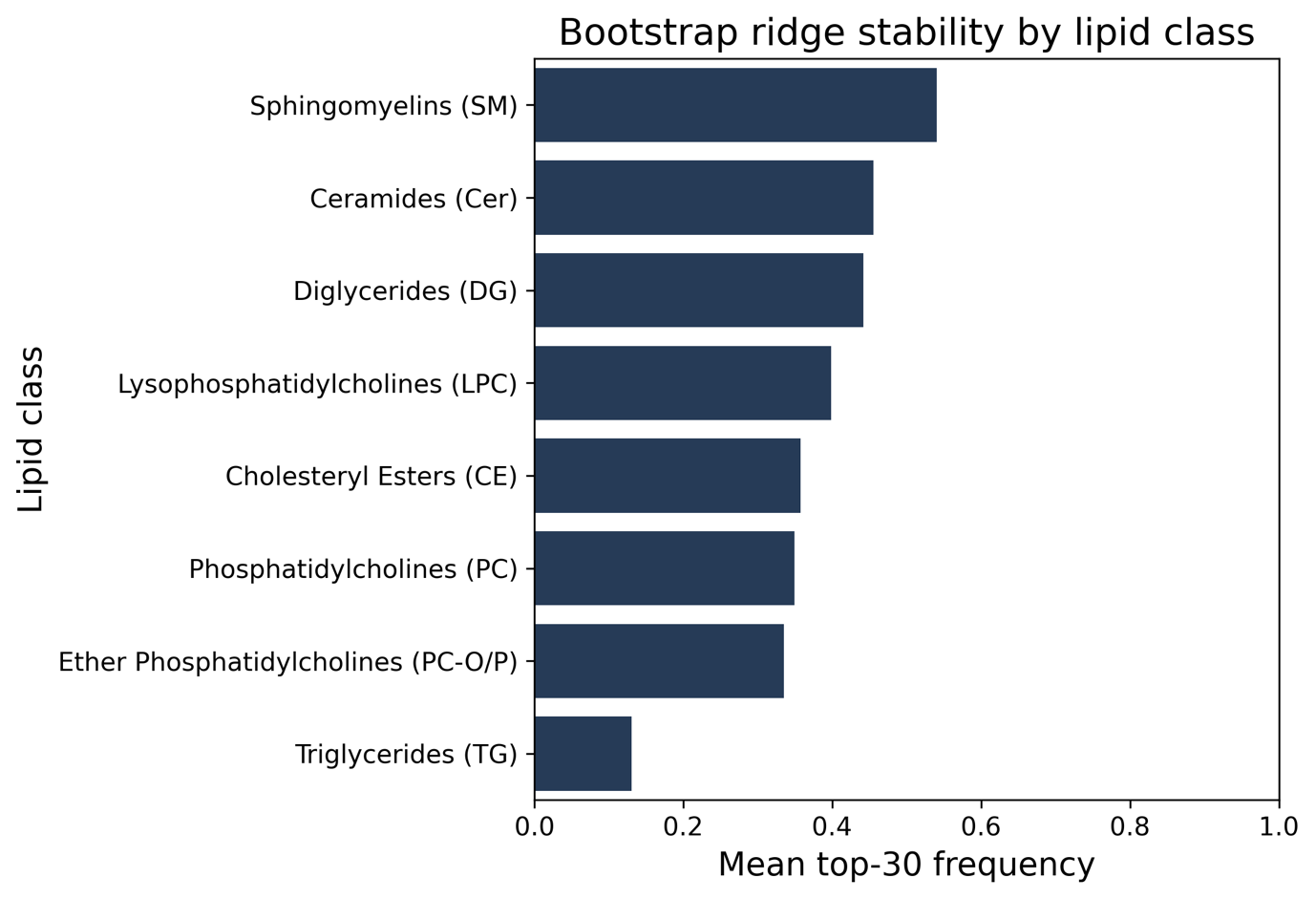

*Sup Fig. 4: Top 30 Frequency by Lipid Class*

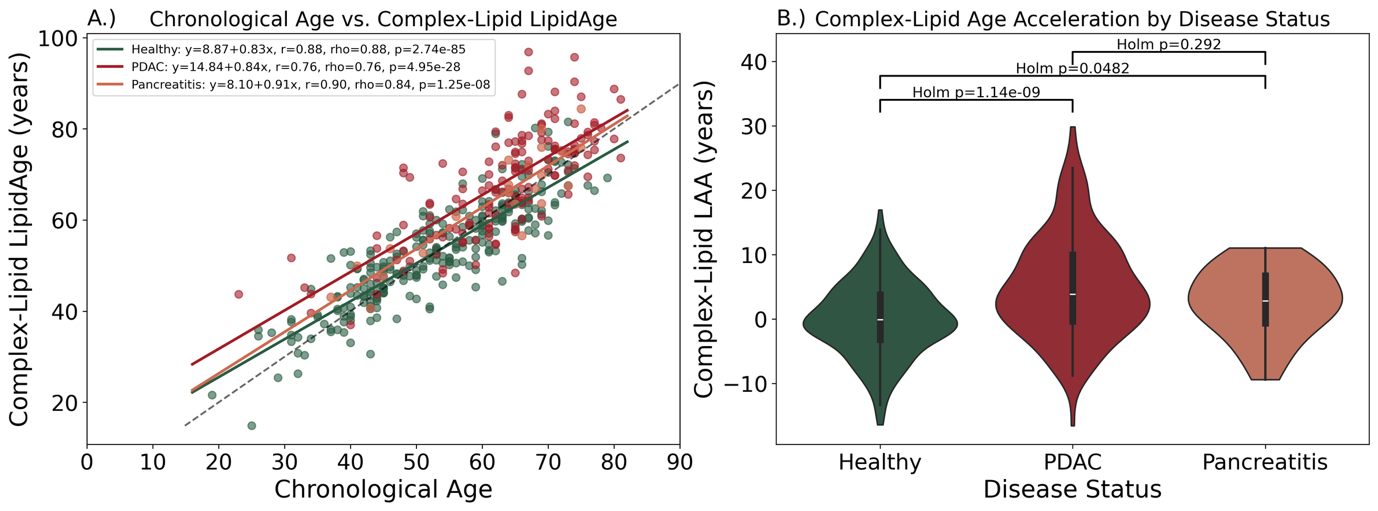

*Sup. Figure 5: Evaluation of the streamlined Complex-Lipid aging clock and corresponding age acceleration across clinical cohorts. A.) Correlation between chronological age and predicted Complex-Lipid LipidAge. Scatter plot demonstrating the linear relationship between chronological age (x-axis) and the biological age predicted using the optimized 55-feature Complex LipidClock framework (y-axis). Regression trajectories and statistical metrics—including Pearson correlation coefficients (r), Spearman’s rank correlation coefficients (ρ), and p-values—are provided for healthy controls (green), PDAC (maroon), and pancreatitis patients (orange). The dashed gray line represents the line of identity (y=x). B.) Quantification of Complex-Lipid Age Acceleration (LAA) by disease status. Violin plots displaying the distribution and density of LAA (measured in years) calculated from the streamlined complex lipid model. Internal box plots represent the median, interquartile range (IQR), and 95% confidence intervals. Group-wise variations were assessed using post-hoc comparisons with Holm-Bonner multiple-testing correction. Highly significant complex lipid age acceleration is observed in PDAC patients relative to healthy controls (p=1.14×10 −9 ). Notably, in this exploratory analysis, pancreatitis patients exhibited statistically significant biological age acceleration relative to healthy individuals (p=0.0482), contrastive to the non-significant trend observed in the full clock model.*

Supplementary Tables

|  | **Mean Absolute Error** | **Median Absolute Error** | **Mean Squared Error** | **Pearson R** |
| --- | --- | --- | --- | --- |
| **Healthy** | 4.82 | 3.46 | 40.31 | 0.86 |
| **PDAC** | 8.09 | 6.80 | 105.77 | 0.71 |
| **Pancreatitis** | 5.18 | 5.08 | 36.48 | 0.88 |

*Sup Tab. 4: Performance Metrics on test data*

| **Metric** | **MDT = 6** | **MDT = 7** | **MDT = 8** | **MDT = 9** | **MDT = 10** |
| --- | --- | --- | --- | --- | --- |
| **OS univariable HR for LAA** | 1.085 | 1.073 | 1.063 | 1.056 | 1.050 |
| **OS univariable p-value** | 1.72E-09 | 1.72E-09 | 1.72E-09 | 1.72E-09 | 1.72E-09 |
| **PFS univariable HR for LAA** | 1.037 | 1.032 | 1.028 | 1.024 | 1.022 |
| **PFS univariable p-value** | 1.88E-02 | 1.88E-02 | 1.88E-02 | 1.88E-02 | 1.88E-02 |
| **OS core-adjusted HR for LAA** | 1.127 | 1.108 | 1.094 | 1.083 | 1.074 |
| **OS core-adjusted p-value** | 3.57E-07 | 3.57E-07 | 3.57E-07 | 3.57E-07 | 3.57E-07 |
| **PFS core-adjusted HR for LAA** | 1.051 | 1.043 | 1.038 | 1.034 | 1.030 |
| **PFS core-adjusted p-value** | 3.30E-02 | 3.30E-02 | 3.30E-02 | 3.30E-02 | 3.30E-02 |

*Sup Tab. 5 :* *Sensitivity of survival associations to the mortality-doubling-time assumption: Sensitivity of univariable and core-adjusted survival associations of continuous lipid age acceleration (LAA) to variation in the mortality-doubling-time (MDT) parameter.*

| **Metric** | **MDT = 6** | **MDT = 7** | **MDT = 8** | **MDT = 9** | **MDT = 10** |
| --- | --- | --- | --- | --- | --- |
| **Healthy Pearson r** | 0.919 | 0.891 | 0.861 | 0.828 | 0.795 |
| **Healthy MAE, years** | 3.620 | 4.220 | 4.820 | 5.420 | 6.030 |
| **PDAC test Pearson r** | 0.823 | 0.771 | 0.718 | 0.665 | 0.614 |
| **PDAC test MAE, years** | 6.070 | 7.090 | 8.100 | 9.110 | 10.120 |
| **Healthy, proportion with LAA > 0** | 0.523 | 0.523 | 0.523 | 0.523 | 0.523 |
| **PDAC test, proportion with LAA > 0** | 0.717 | 0.717 | 0.717 | 0.717 | 0.717 |
| **Pancreatitis, proportion with LAA > 0** | 0.727 | 0.727 | 0.727 | 0.727 | 0.727 |

*Sup Tab. 6:* *Sensitivity of age-prediction performance to the mortality-doubling-time assumption: Sensitivity of age-correlation, absolute error, and LAA-positive fractions to variation in the mortality-doubling-time (MDT) parameter*

| **Covariate** | **Coefficient (β)** | **Standard Error (SE)** | **Hazard Ratio (HR)** | **95% Confidence Interval (CI)** | **z-statistic** | **p-value** |
| --- | --- | --- | --- | --- | --- | --- |
| **Lipid Age Acceleration (LAA)** | 0.085 | 0.018 | 1.09 | [1.05, 1.13] | 4.76 | 2.18×10−6 |
| **Chronological Age** | 0.046 | 0.012 | 1.05 | [1.02, 1.07] | 3.85 | 1.17×10−4 |
| **Sex (Male vs. Female)** | 0.149 | 0.238 | 1.16 | [0.73, 1.85] | 0.62 | 0.532 |
| **Body Mass Index (BMI)** | 0.002 | 0.018 | 1 | [0.97, 1.04] | 0.14 | 0.891 |
| **Diabetes Mellitus (Yes)** | 0.529 | 0.26 | 1.7 | [1.02, 2.82] | 2.04 | 0.042 |
| **Treatment Status (Treated)** | -1.121 | 0.413 | 0.33 | [0.14, 0.73] | -2.71 | 0.007 |

*Sup Tab. 7 : Adjusted model for treatment status*

| **Endpoint & Model Specification** | **Patients (N)** | **Events (n)** | **LAA Hazard Ratio (95% CI)** | **P-value** | **C-index** |
| --- | --- | --- | --- | --- | --- |
| **Progression Free Survival (PFS)** |  |  |  |  |  |
| **Model 1: LipidAgeAcceleration** | 145 | 104 | 1.03 (1.00-1.05) | <0.05 | 0.61 |
| **Model 1.1: Crude Model: LipidAgeAcceleration (cohort matched)** | 85 | 63 | 1.02 (0.99-1.05) | 0.13 | 0.61 |
| **Model 2: Core Clinical Adjustments(LAA + Age + sex + BMI + diabetes)** | 85 | 63 | 1.04 (1.00-1.07) | <0.05 | 0.66 |
| **Model 3: Core + CA19-9** | 85 | 63 | 1.02 (0.98-1.06) | 0.31 | 0.70 |
| **Model 4: Core + Metastasis (M-status)** | 70 | 56 | 1.07 (1.02-1.12) | <0.005 | 0.71 |
| **Model 5:Core + M + G** | 48 | 38 | 1.05 (0.99-1.11) | 0.14 | 0.71 |
| **Model 6: Core + TNM** | 47 | 38 | 1.02(0.96-1.09) | 0.53 | 0.73 |
| **Model 7: Core + TNMG Tumour Burden (Complete-Case)** | 37 | 29 | 1.01 (0.93-1.09) | 0.84 | 0.85 |
| **Model 8: Core + TNMG Tumor Burden (Missing-Indicator)** | 85 | 63 | 1.03 (0.98-1.07) | 0.22 | 0.73 |

*Sup Tab. 8: Multivariable Cox proportional hazards regression models for Progression Free Survival (PFS) based on Lipid Age Acceleration (LAA).*

| **Covariate** | **Hazard Ratio** | **95% CI** | **SE** | **Z** | **p-value** |
| --- | --- | --- | --- | --- | --- |
| **TG 58:2** | 1.345 | 1.035-1.748 | 0.134 | 2.217 | 0.027 |
| **SM 41:2** | 0.61 | 0.407-0.934 | 0.212 | -2.282 | 0.022 |

*Sup Tab. 9:* *Cox Proportional Hazard model for lipidome and covariates with coefficient and significant HR*

2. Wolrab, D. et al. Validation of lipidomic analysis of human plasma and serum

by supercritical fluid chromatography-mass spectrometry and hydrophilic

interaction liquid chromatography-mass spectrometry. *Anal. Bioanal. Chem.* 2020;412(10):2375. doi: 10.1007/s00216-020-02473-3

3.	Triebl A, Burla B, Selvalatchmanan J, et al. Shared reference materials harmonize lipidomics across MS-based detection platforms and laboratories. *J Lipid Res*. 2020;61(1):105-115. doi:10.1194/jlr.D119000393

3.	Fong S, Pabis K, Latumalea D, et al. The principal component-based clinical aging clock (PCAge) identifies signatures of healthy aging and provides normative targets for clinical intervention. Published online July 16, 2023:2023.07.14.23292604. doi:10.1101/2023.07.14.23292604

5.	Gompertz B. XXIV. On the nature of the function expressive of the law of human mortality, and on a new mode of determining the value of life contingencies. In a letter to Francis Baily, Esq. F. R. S. &c. *Philosophical Transactions of the Royal Society of London*. 1825;115:513-583. doi:10.1098/rstl.1825.0026

6.	Davidson-Pilon C. lifelines: survival analysis in Python. *Journal of Open Source Software*. 2019;4(40):1317. doi:10.21105/joss.01317
